# Decline in Emergent and Urgent Care during the COVID-19 Pandemic

**DOI:** 10.1101/2020.05.14.20096602

**Authors:** Dhruv S. Kazi, Rishi K. Wadhera, Changyu Shen, Kalon K. L. Ho, Rushad Patell, Magdy H. Selim, John Urwin, Mark L. Zeidel, Peter Zimetbaum, Kevin Tabb, Robert W. Yeh

**Affiliations:** Richard A. and Susan F. Smith Center for Outcomes Research in Cardiology, Division of Cardiology, Beth Israel Deaconess Medical Center, Boston, Massachusetts; Department of Medicine, Beth Israel Deaconess Medical Center, Boston, Massachusetts; Harvard Medical School, Boston, Massachusetts; Division of Hematology, Beth Israel Deaconess Medical Center, Boston, Massachusetts; Department of Neurology, Beth Israel Deaconess Medical Center, Boston, Massachusetts; Beth Israel Lahey Health, Cambridge, Massachusetts

## Abstract

Due to the ongoing coronavirus disease (COVID-19) pandemic, there are concerns that patients may be avoiding care for emergent and urgent health conditions due to fear of contagion or as an unintentional consequence of government orders to postpone “non-essential” services. We therefore sought to evaluate the effect of the COVID-19 pandemic on the number of patient encounters for select emergent or urgent diagnoses at a large tertiary-care academic medical center in Boston. Inpatient diagnoses included acute myocardial infarction (MI) and stroke, and outpatient but urgent diagnoses included new referrals for breast and hematologic malignancies. For each condition, we used a “difference-in-differences” approach to estimate the proportional change in number of encounters during the pandemic (March – April 2020) compared with earlier in the same year (January – February 2020), using equivalent periods in 2019 as a control. After the onset of the pandemic, we observed significant reductions in hospitalizations for MI (difference-in-differences estimate, 0.67; 95%CI, 0.46-0.96; P=0.04) and stroke (difference-in-differences estimate, 0.42; 95%CI, 0.28-0.65; P<0.001) (Table). In the ambulatory setting, there was a reduction in referrals for breast cancer and hematologic cancers, but this did not reach statistical significance until the month after the onset of the pandemic. Our findings suggest an urgent need for public health messaging to ensure that patients continue to seek care for acute emergencies. In addition, decisions by health systems regarding when to reinitiate non-emergent care should carefully factor in the harms of delayed diagnosis and treatment occurring during the COVID-19 pandemic.

## MANUSCRIPT

### Background

In early March 2020, the number of confirmed cases of novel coronavirus disease (COVID-19) in Massachusetts began increasing rapidly. In response to the spread, Massachusetts declared a state of emergency on March 10, 2020. Five days later, the Massachusetts Department of Public Health directed all hospitals and ambulatory surgical centers to postpone or cancel any nonessential procedures to focus the use of healthcare resources on pandemic response and conserve personal protective equipment.^1^ Although life-sustaining interventions are explicitly excluded from the order, patients may not be able to distinguish between elective and urgent or emergent problems, and may defer necessary care due to fear of contagion.

### Objective

We therefore sought to evaluate the effect of the COVID-19 pandemic on the number of patient encounters for select emergent or urgent diagnoses at the Beth Israel Deaconess Medical Center, a large tertiary-care academic medical center in Boston.

### Methods and Findings

We selected conditions for which all patients would be expected to seek care under normal circumstances. Inpatient diagnoses included acute myocardial infarction (MI) and stroke, and outpatient but urgent diagnoses included new referrals for breast and hematologic malignancies. We included breast cancer as it is a common malignancy diagnosed by screening procedures and hematologic cancers as they typically present with mild symptoms yet require urgent treatment. For each condition, we used a “difference-in-differences” approach to estimate the proportional change in number of encounters during the pandemic (March – April 2020) compared with earlier in the same year (January – February 2020), using equivalent periods in 2019 as a control. Because the effect on referrals for new malignancies may be delayed (as they would be expected to occur after a decline in primary care clinic visits and screening tests), we performed a sensitivity analysis including a 1-month lag for the oncology referrals. As these analyses used de-identified count data collected as part of a systemwide quality improvement initiative, the institutional review board at Beth Israel Deaconess Medical Center deemed the project not to constitute human subjects research.

After the onset of the pandemic, we observed significant reductions in hospitalizations for MI (difference-in-differences estimate, 0.67; 95%CI, 0.46-0.96; P=0.04) and stroke (difference-in-differences estimate, 0.42; 95%CI, 0.28-0.65; P<0.001) (Table). In the ambulatory setting, there was a numerical decline in new oncology referrals starting March 1, 2020, but these did not achieve statistical significance in the base-case analysis. However, in lag analyses that examined the effect on cancer diagnoses starting April 1, 2020, there was a significant reduction in referrals for breast cancer (difference-in-differences estimate, 0.35; 95%CI, 0.21-0.58; P<0.001) and hematologic cancers (difference-in-differences estimate, 0.39; 95% CI, 0.22-0.68; P<0.001).

### Discussion

We found a marked decline in presentations for emergent cardiovascular conditions during the pandemic. Reductions in MI during the pandemic have also been observed in Italy,^2^ and for ST-elevation MI in the US,^3^ but reductions in stroke have not been described before. This decline may be partially due to a true reduction in incident events due to factors such as altered lifestyle and improved air quality during the lockdown. However, Massachusetts has experienced a large increase in at-home mortality during the COVID-19 pandemic, raising concern that people may be deferring necessary care out of fear of contagion, and possibly dying at home.^4^ Furthermore, short-term lifestyle changes cannot explain the decline in referrals for new diagnoses of malignancies observed in our study, which are likely the result of decreased outpatient cancer screening during this period, due to postponement of in-person primary care visits and screening studies, and delayed referrals.^5^ Given the time-sensitive nature of initiating treatment for these and other life-threatening conditions, urgent efforts are needed to improve public health messaging to ensure that patients continue to seek care for acute emergencies. In addition, decisions by health systems regarding when to reinitiate non-emergent care should carefully factor in the harms of delayed diagnosis and treatment occurring during the COVID-19 pandemic.

## Data Availability

All relevant data are included in the manuscript.

## COMPETING INTERESTS

Authors have disclosed no conflicts of interest.

## AUTHOR APPROVAL

All authors have seen and approved the manuscript prior to submission.

## AUTHOR CONTRIBUTIONS

Conception and design: D. Kazi, R. Wadhera, K. Tabb, R. Yeh.

Drafting of the article: D. Kazi.

Critical revision of the article for important intellectual content: D. Kazi, R. Wadhera, C. Shen, K. Ho, R. Patell, M Selim, J Urwin, M Zeidel, P Zimetbaum, K Tabb, R Yeh.

Final approval of the article: D. Kazi, R. Wadhera, C. Shen, K. Ho, R. Patell, M Selim, J Urwin, M Zeidel, P Zimetbaum, K Tabb, R Yeh.

Administrative, technical, or logistic support: D. Kazi.

## PRIMARY FUNDING SOURCE

None.

## DATA AVAILABILITY STATEMENT

All encounter data used in the analyses are presented in the Table.

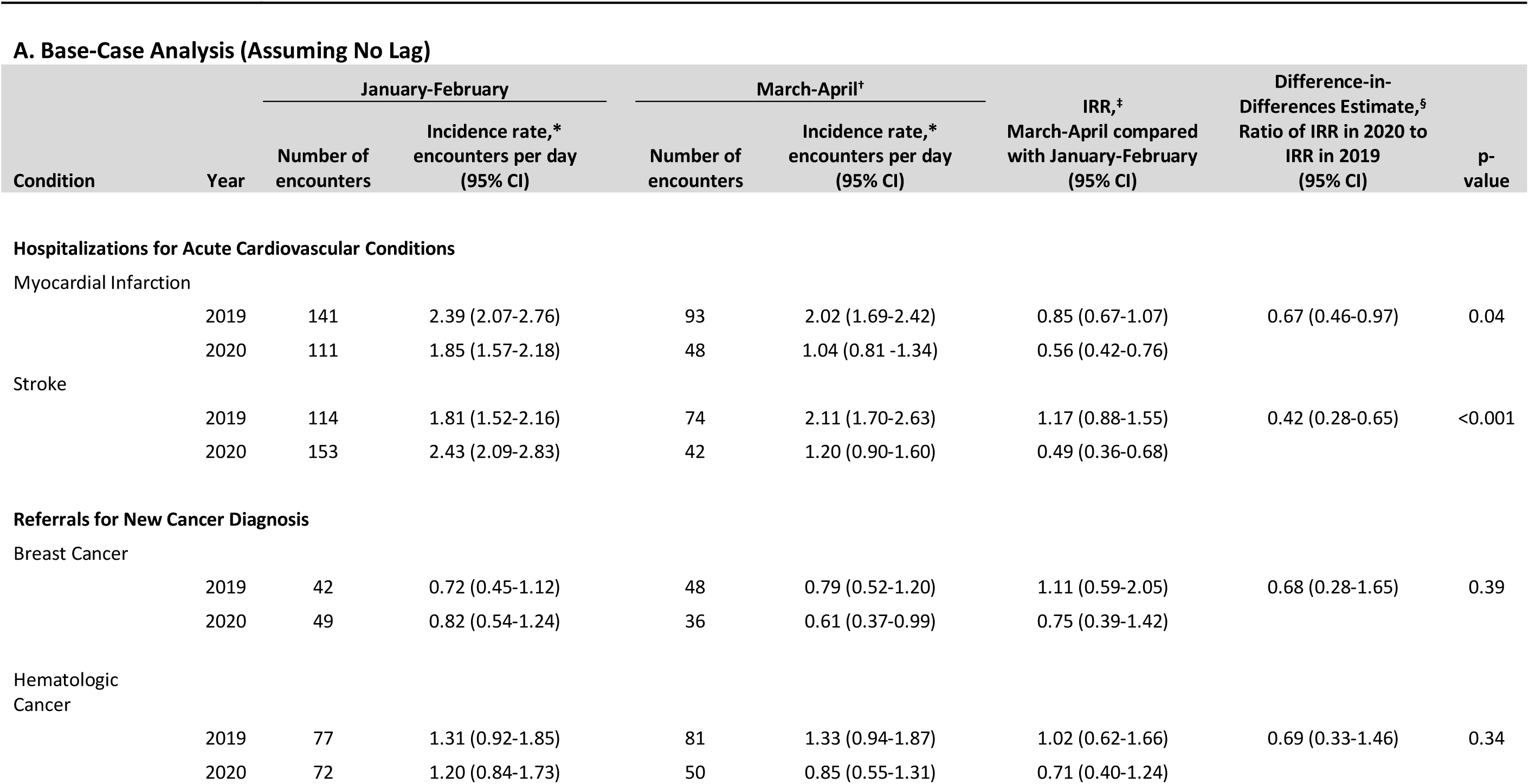
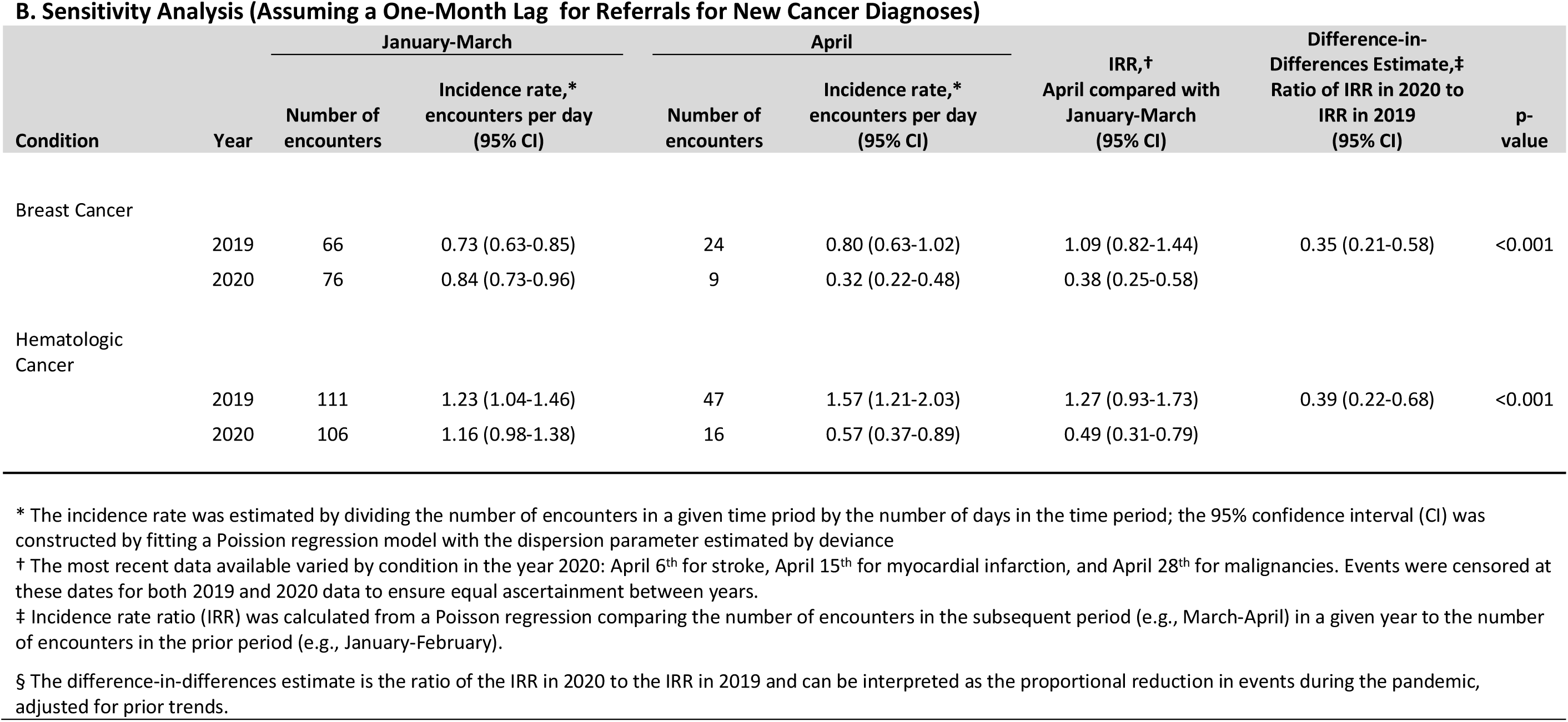
**Table. Results**. Changes in Encounters per Day Associated with the COVID-19 Pandemic

## Notes

### Competing Interest Statement

The authors have declared no competing interest.

### Clinical Trial

N/A

### Funding Statement

No external funding for this work.

